# Vision and Language Models for Classifying Maxillary Sinus Disease on Cone-Beam Computed Tomography: A Transparent Multimodal Benchmark

**DOI:** 10.64898/2026.08.11.26360189

**Authors:** Seba Al-Hebshi, Hanadi Khalifa, Tuan D. Pham

## Abstract

**Background:** Cone-beam computed tomography (CBCT) frequently captures the maxillary sinuses incidentally, and reliable automated detection of sinus abnormality is clinically relevant. Unlike most vision–language benchmarks in medical imaging, which pair images with pre-existing, human-authored clinical reports, findings text can also be generated directly by a large language model from the image itself—raising the question of how much diagnostic value such AI-derived text carries, and whether that value depends on independent verification. Multimodal artificial intelligence (AI) benchmarks risk overstating performance if the provenance of each input—image, raw AI-generated text, or radiologist-verified text—is not clearly separated and reported.

**Methods:** We used 300 mid-sagittal CBCT slices from the MMDental dataset. ChatGPT generated findings text and a provisional normal/abnormal label for every slice (majority vote, three independent readings from the image alone); primary classification performance was assessed on this full, unfiltered set (n=300). A radiologist then independently reviewed each case’s image together with ChatGPT’s description, producing their own diagnosis; three cases were excluded as insufficient, yielding 297 confirmed cases. On this subset, every model was retrained and re-evaluated under identical 10-fold cross-validation on both the provisional ChatGPT-only labels (“pre”) and the radiologist-confirmed labels (“post”), isolating the effect of label provenance from image or architecture. Eight vision architectures, seven language classifiers, and five VLMs were evaluated throughout; three generative models performed exploratory note-drafting.

**Findings:** Raw ChatGPT-generated text produced the highest performance of any modality or condition: language models reached near-ceiling AUC (0·992 to 1·000, n=300), exceeding every vision model (AUC 0·799 to 0·880) and every VLM image-only probe (AUC 0·63 to 0·69). On the 297-case pre/post analysis, this advantage depended heavily on label source: language and text-derived VLM performance fell substantially from ChatGPT-only to radiologist-confirmed labels (e.g. BERT-base AUC 0·999 to 0·837), while vision-model performance was stable or modestly improved (e.g. DenseNet-121 0·867 to 0·891). The radiologist reclassified 62 of 297 cases (21%) relative to ChatGPT’s provisional read, and a meaningful proportion of raw ChatGPT text was clinically uninterpretable or unsupported by the imaging.

**Interpretation:** As shown here for the first time, raw, image-derived AI-generated text yields the highest apparent classification performance in this benchmark, but this reflects the text’s alignment with its own self-generated labels rather than verified diagnostic content, and a substantial share of that text is not clinically explainable. Radiologist-confirmed text and labels give a lower but trustworthy estimate of true performance, on which convolutional neural network (CNN) vision models remain a stable, comparatively inexpensive baseline. Multimodal dental AI should report performance separately by modality and label provenance rather than pooling headline metrics.

**Research in context:** *Evidence before this study:* We searched PubMed with the terms “maxillary sinus”, “CBCT”, “deep learning”, “artificial intelligence”, and “vision–language model” from January 2015 to July 2026. Published benchmarks have predominantly evaluated single-modality convolutional networks for dental CBCT tasks, with limited transparency about how AI-generated text inputs are produced or verified. No study had systematically compared image-only, language-only, and VLM families for maxillary sinus classification on the same dataset under matched cross-validation, nor had the extent to which apparent multimodal performance depends on whether AI-generated text is used raw or independently verified been explicitly characterised.

*Added value of this study:* Unlike prior vision–language benchmarks in medical imaging, which typically pair images with pre-existing, human-authored clinical reports, this study evaluates text generated directly by an LLM from the image itself, independent of any prior radiological report. This design isolates a question distinct from conventional report-grounded VLM evaluation: not how well a model can classify existing clinical documentation, but how much apparent diagnostic value an LLM’s own image-derived description carries—and how much of that apparent value depends on whether the description is used raw or independently verified. We show that raw, unverified ChatGPT-generated text produces the highest classification performance of any modality evaluated (AUC up to 1·000), exceeding image-only and image-plus-text models—but that a meaningful proportion of this raw text is clinically uninterpretable, and that its apparent advantage collapses substantially once evaluated against a radiologist-confirmed reference standard on the same cases. Trained vision models, by contrast, remain stable regardless of label source, indicating that their signal derives from image content rather than label provenance. We further show that ViTs underperform CNNs at dental-imaging dataset scale.

*Implications of all the available evidence:* Raw AI-generated text can substantially outperform image-based classification on headline accuracy metrics, but this performance is not synonymous with clinical trustworthiness or explainability and should not be reported or deployed without independent verification. CNNs remain reliable, comparatively low-cost baselines for sinus CBCT screening at small dataset sizes. Multimodal dental AI benchmarks must report modality-specific results transparently and distinguish raw from verified text inputs; headline metrics should not be pooled across these conditions without disclosing their provenance. Generative VLMs show near-term utility as documentation aids rather than autonomous diagnostic systems, and full three-dimensional CBCT analysis remains the essential next step for clinical translation in the maxillofacial trauma setting.

## Introduction

Cone-beam computed tomography (CBCT) is widely used in dentistry for implant planning, endodontics, and oral and maxillofacial surgery, and frequently captures the maxillary sinuses within its field of view. Incidental sinus findings—most commonly mucosal thickening, retention pseudocysts, and partial opacification—are detected in a substantial proportion of dental CBCT examinations and can influence treatment planning, particularly before sinus-floor elevation and posterior implant placement.^1,2^ Reliable, reproducible identification of an abnormal maxillary sinus on CBCT is therefore clinically relevant, yet reporting is operator-dependent and time-consuming.

Deep learning has been applied increasingly to dental and maxillofacial imaging, where convolutional neural networks (CNNs) can support detection, classification, and triage.^3^ More recently, the field has expanded to include language models and multimodal vision–language models (VLMs), most of which are evaluated by pairing images with pre-existing, human-authored clinical reports. General-purpose large language models (LLMs) such as ChatGPT raise a different possibility: rather than classifying an existing report, an LLM can generate its own structured radiological description directly from the image, with no prior human report as input. This is conceptually distinct from conventional report-grounded VLM evaluation, and raises a question that has not been systematically addressed: how much diagnostic value does an LLM’s own image-derived text carry, and does that value depend on whether the text is used as generated or independently verified by a clinician?

Prior work has demonstrated the clinical potential of deep learning and vision–language approaches for paediatric dental disease diagnosis using panoramic radiographs,^4,5^ establishing that effective AI-assisted dental diagnosis is achievable with publicly available architectures and without proprietary imaging infrastructure. The present study extends that research programme to adult maxillofacial imaging and, for the first time, formally evaluates image-derived ChatGPT text—both raw and radiologist-verified—as a diagnostic classifier, benchmarked alongside eight vision architectures, seven language models, and five VLMs under matched cross-validation.

Such comparisons require care in interpretation. Text generated by an LLM to describe an image it has itself classified can let language models attain exceptionally high scores that reflect agreement with the LLM’s own self-generated label rather than independent, verifiable image interpretation. General-purpose VLMs have rarely been validated on niche modalities such as sinus CBCT and may default to a single class. Vision Transformers (ViTs), although state of the art on large natural-image corpora, are well-known data-hungry and may underperform CNNs on the small datasets typical of dental research. A benchmark that reports a single headline metric per model, without distinguishing raw AI-generated text from independently verified text, risks presenting an artefact of label self-consistency as a genuine diagnostic advance.

In this study we report a deliberately transparent multimodal benchmark for normal-versus-abnormal maxillary sinus classification on a large public CBCT dataset. Our objectives were fourfold: (i) to establish an image-only baseline across eight modern vision architectures under identical cross-validation; (ii) to determine how much diagnostic performance is carried by raw, image-derived ChatGPT text, evaluated on the full unfiltered dataset; (iii) to test how that performance changes when the same models are retrained and re-evaluated against an independent radiologist-confirmed reference standard, isolating the effect of verification from image or architecture; and (iv) to evaluate contrastive and generative VLMs under matched linear-probe cross-validation, including an exploratory use of generative VLMs for AI-assisted note drafting. The contribution is a demonstration that image-derived AI-generated text can outperform image-based classification on raw metrics, together with a transparent account of how much of that advantage depends on independent clinical verification.

## Methods

### Study dataset and reference standard

We used the publicly available MMDental dataset,^6^ from which 300 mid-sagittal maxillary-sinus CBCT slices were drawn. Examples of ChatGPT’s raw descriptions alongside the radiologist’s corrections are given in Tables 1 and 2. Every classification model was evaluated under three conditions, distinguished by data source and label provenance:

1. Raw (n=300): the full, unfiltered dataset, using ChatGPT’s own image-derived text and labels throughout.
2. Pre (n=297; 146 normal, 151 abnormal) using ChatGPT’s provisional descriptions.
3. Post (n=297; 146 normal, 151 abnormal): the same 297 cases, using the radiologist-confirmed reference standard.

**Table 1.** ChatGPT and radiologist descriptions for 10 representative maxillary-sinus CBCT cases (five classified as normal, rows 1–10; five as abnormal, rows 11–20, by ChatGPT). Each case is shown as a ChatGPT row followed by a Radiologist row giving the radiologist’s classification and diagnosis; three of the ten cases were reclassified relative to the ChatGPT-only label.

| Image | Normal | Description |
| --- | --- | --- |
| 9.1 | ChatGPT | Well-aerated maxillary sinus, preserved sinus cavity, no mucosal thickening, no retention pseudocyst, no air-fluid level, no opacification, intact sinus walls. |
|  | Radiologist | Classification: Normal. Diagnosis: Normal / no significant finding. |
| 214.7 | ChatGPT | Well-aerated maxillary sinus, no appreciable mucosal thickening, no focal soft-tissue opacity, no air-fluid level, no cortical destruction or expansion. |
|  | Radiologist | Classification: Abnormal. Diagnosis: Mucosal thickening (Mild (<3mm)). |
| 212.6 | ChatGPT | Well-aerated maxillary sinus, no appreciable mucosal thickening, no focal soft-tissue lesion, no air-fluid level, no cortical destruction or expansion. |
|  | Radiologist | Classification: Normal. Diagnosis: Normal / no significant finding. |
| 205.6 | ChatGPT | Well-aerated maxillary sinus, no mucosal thickening, no opacification, no air-fluid level, no cortical destruction or expansion. |
|  | Radiologist | Classification: Normal. Diagnosis: Normal / no significant finding. |
| 416.12 | ChatGPT | Preserved maxillary sinus aeration, no significant mucosal thickening, no air-fluid level, no opacification, no cortical destruction. |
|  | Radiologist | Classification: Abnormal. Diagnosis: Mucosal thickening (Mild (<3mm)). |
| **Abnormal** |  |  |
| 295.8 | ChatGPT | Dome-shaped lesion arising from sinus floor, well-circumscribed soft tissue opacity, preserved surrounding aeration, no air-fluid level, no cortical destruction. |
|  | Radiologist | Classification: Abnormal. Diagnosis: Retention cyst / pseudocyst. |
| 4.4 | ChatGPT | Near-complete maxillary sinus opacification, severe loss of sinus aeration, no discrete retention pseudocyst identified, no air-fluid level, intact sinus walls, no cortical destruction or expansion. |
|  | Radiologist | Classification: Abnormal. Diagnosis: Sinus opacification - complete (>90%). |
| 342.11 | ChatGPT | Dome-shaped soft tissue opacity, lesion arising from sinus floor, partial inferior sinus occupancy, preserved superior aeration, no air-fluid level, no cortical destruction. |
|  | Radiologist | Classification: Abnormal. Diagnosis: Retention cyst / pseudocyst. |
| 425.13 | ChatGPT | Unilateral maxillary sinus mucosal thickening, inferior sinus floor involvement, mild-to-moderate extent, preserved overall aeration, no air-fluid level, no cortical destruction or expansion. |
|  | Radiologist | Classification: Normal. Diagnosis: Normal / no significant finding. |
| 376.7 | ChatGPT | Horizontal air-fluid level in maxillary sinus, partial inferior sinus opacification, preserved superior aeration, no cortical destruction, possible odontogenic relationship. |
|  | Radiologist | Classification: Abnormal. Diagnosis: Mucosal thickening (Moderate (3-6mm)). |

**Table 2.** Representative cases among the 62 normal/abnormal reclassifications between the provisional ChatGPT-only label and the radiologist-confirmed reference standard. For each case the table gives ChatGPT’s pre-correction classification and description alongside the radiologist’s post-correction classification and diagnosis; cases were selected where the radiologist’s correction added the most diagnostic detail, rather than a bare label change.

| Image | Source | Description |
| --- | --- | --- |
| 477.17 | ChatGPT (pre) | Classification: Normal. Well-aerated maxillary sinus, no mucosal thickening, no sinus opacification, no retention pseudocyst, no polypoid lesion, no air-fluid level, intact sinus walls, no cortical destruction, no expansion. |
|  | Radiologist (post) | Classification: Abnormal. Diagnosis: Mucosal thickening (Moderate (3–6mm)). |
| 239.10 | ChatGPT (pre) | Classification: Normal. Well-aerated maxillary sinus, preserved sinus cavity, no mucosal thickening, no retention pseudocyst, no polypoid lesion, no opacification, no air-fluid level, intact sinus walls. |
|  | Radiologist (post) | Classification: Abnormal. Diagnosis: Retention cyst / pseudocyst. |
| 203.12 | ChatGPT (pre) | Classification: Normal. Well-aerated maxillary sinus, preserved sinus cavity, no mucosal thickening, no retention pseudocyst, no air-fluid level, no opacification, intact sinus walls. |
|  | Radiologist (post) | Classification: Abnormal. Diagnosis: Mucosal thickening (Mild (<3mm)). |
| 364.6 | ChatGPT (pre) | Classification: Abnormal. Unilateral dome-shaped lesion arising from the maxillary sinus floor, focal soft-tissue opacity, preserved sinus aeration, no air-fluid level, no cortical destruction, no sinus expansion. |
|  | Radiologist (post) | Classification: Normal. Diagnosis: Normal / no significant finding. |
| 4.6 | ChatGPT (pre) | Classification: Abnormal. Irregular inferior maxillary sinus mucosal thickening, polypoid component, preserved superior aeration, no air-fluid level, no cortical destruction, no sinus expansion, limited single-slice assessment. |
|  | Radiologist (post) | Classification: Normal. Diagnosis: Normal / no significant finding. |

Conditions 2 and 3 share an identical 10-fold stratified cross-validation split and case-to-fold assignment, isolating the effect of label provenance from image content or model architecture; condition 1 establishes the headline, unfiltered performance ceiling against which conditions 2 and 3 are then interpreted. “Abnormal” encompassed mucosal thickening, retention pseudocyst, opacification, and air–fluid level; “normal” denoted a well-aerated sinus with intact walls. The overall study design and analysis pipeline is summarised in Figure 1.

**Figure 1.**
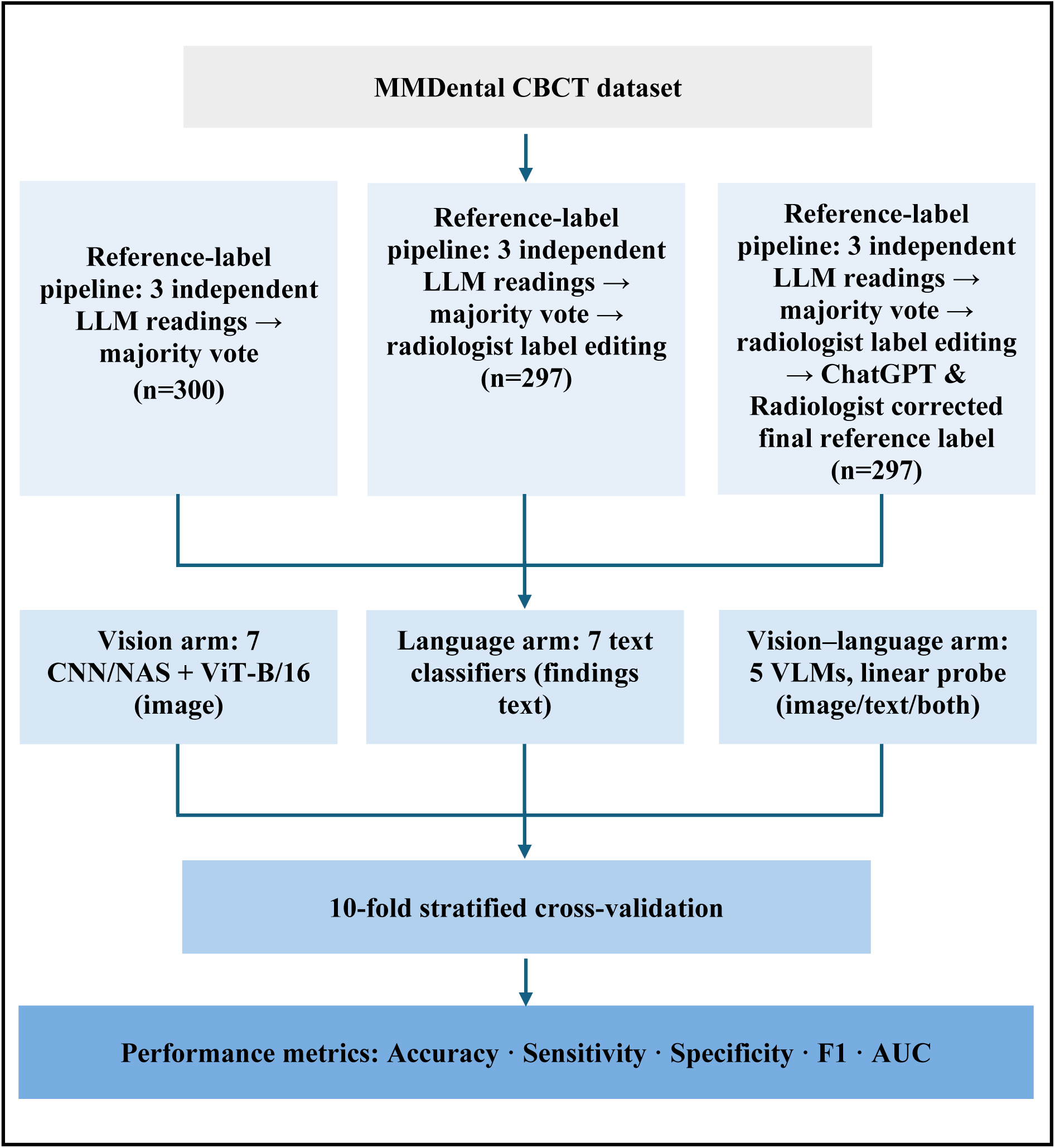
Study design and analysis pipeline. The dataset passed through a three-stage reference-label pipeline (LLM majority vote followed by radiologist confirmation) before being distributed to the three model arms, each evaluated under matched 10-fold stratified cross-validation.

### Ethics

This study is a secondary analysis of the publicly available, fully de-identified MMDental dataset. Original data collection, informed consent, and anonymisation were approved by the Medical Ethics Committee of Lishui University (approval identification number: 2022YR014), in accordance with the principles of the Declaration of Helsinki, and the committee separately approved open publication of the anonymised dataset under a Creative Commons Attribution (CC-BY) licence. No additional institutional ethics approval was required for this secondary analysis, as no identifiable patient information was accessed at any stage.

### Text generation

For every case, ChatGPT was prompted three times, independently, from the CBCT image alone, with the following instruction: “Act as a consultant oral and maxillofacial radiologist. Analyse this CBCT image focusing ONLY on the paranasal sinus region visible in the scan. 1. identify the MOST likely radiographic description from the following categories when applicable: mucosal thickening - sinusitis - retention pseudocyst - complete/partial opacification - polypoid lesion - air-fluid level - antrolith - oroantral communication - mucocele - fungal sinusitis - post-surgical change - etc. 2. Describe: laterality (right/left/bilateral) - location within sinus - extent/severity (mild/moderate/severe) - relationship to adjacent teeth if relevant - any cortical destruction or expansion - presence/absence of fluid level”. The three resulting analyses for each case were compared, and a majority vote based on similarity across all three readings was used to select a representative ChatGPT-generated description and its associated normal-versus-abnormal classification; this constitutes the raw, unfiltered dataset (n=300; condition 1 above). The model’s structured output comprised this classification, a short description, a confidence rating, and key imaging findings.

This ChatGPT-generated description, together with the corresponding CBCT image, was then given to a radiologist, who independently reviewed both and produced their own diagnosis based on their clinical and scientific expertise; this review was additionally checked by a prosthodontist member of the research team. Three cases were excluded as insufficient, yielding 297 confirmed cases and establishing the radiologist-confirmed reference standard used in the pre/post analysis (conditions 2 and 3 above). For language-model experiments the explicit classification line was removed from every description and only the de-labelled key-findings text was retained as model input. The 10 worked examples in Table 1 are drawn verbatim from these generated descriptions.

### Image preprocessing

All slices were resized to 224×224 pixels, converted from single-channel greyscale to three channels, and normalised using ImageNet statistics. Training-set augmentation comprised horizontal flipping, rotation up to ±15°, mild shift/scale, and small brightness/contrast jitter, applied on the fly; validation images received resizing and normalisation only. Augmentation was applied only during training folds of seven CNNs and a ViT (ViT-B/16); it was not applied to VLMs whose encoders were frozen.

### Vision models

Eight imaging architectures were evaluated: five convolutional networks, ResNet-50,^7^ DenseNet-121,^8^ EfficientNet-B0,^9^ GoogLeNet/Inception,^10^ and AlexNet^11^, two neural architecture search (NAS) networks, NASNet-Large^12^ and PNASNet-5-Large^13^, and ViT-B/16.^14^ All were initialised from ImageNet-pretrained weights and fine-tuned for the binary task.

### Language models

Seven text classifiers were trained on the de-labelled “key findings” field: two models with embeddings learned from scratch (a bidirectional long short-term memory (LSTM) network and a text CNN) and five transformers, BERT-base,^15^ RoBERTa-base,^16^ DistilBERT,^17^ BioBERT,^18^ and Bio_ClinicalBERT.^19^ The explicit “Classification: normal/abnormal” line was removed from every input; only the radiological findings phrases were retained.

### Generative note drafting

Three generative VLMs, Qwen2·5-VL-7B,^20^ MedGemma-4B,^21^ and LLaVA-Med v1·5^22^, were used in an exploratory image-plus-findings note-drafting task. Each model received the CBCT slice together with the de-labelled findings and was prompted to produce a structured Findings/Impression note. Representative outputs for one normal and one abnormal case are shown in Figure 2. This task is framed strictly as AI-assisted documentation, not autonomous diagnosis.

**Figure 2.**
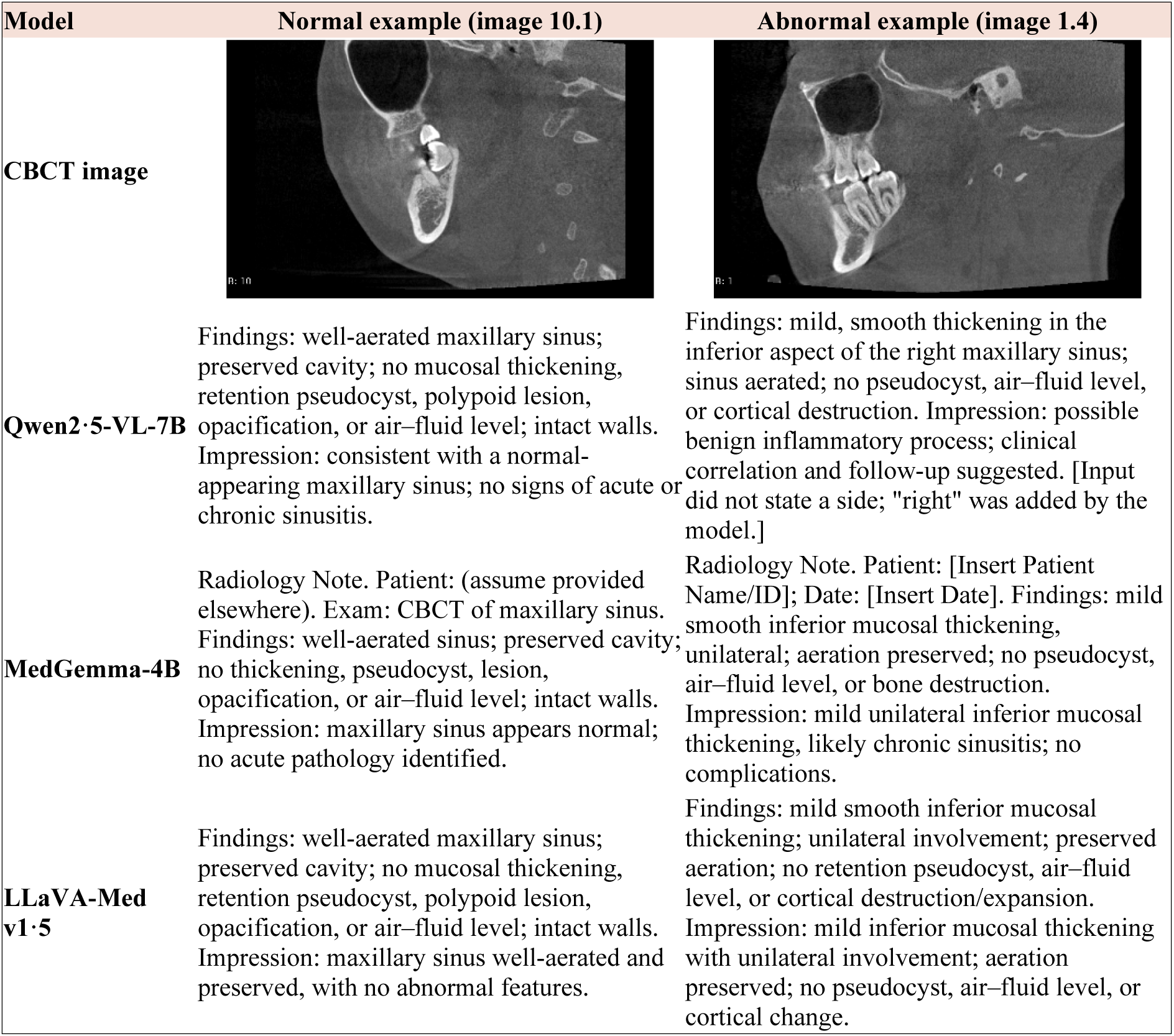
Representative AI-assisted notes generated by the three VLMs for one normal (image 10.1) and one abnormal (image 1.4) CBCT case. Each model received the image together with the de-labelled key findings as input and was prompted to produce a structured Findings/Impression note. Notes are AI-assisted documentation, not independent image diagnosis.

### VLMs under linear probing

Five VLMs, CLIP ViT-B/16,^23^ BiomedCLIP,^24^ Qwen2·5-VL, MedGemma-4B, and LLaVA-Med v1·5, were evaluated as frozen feature extractors under a linear-probe protocol. Pretrained encoder weights were kept frozen; each case was passed through the network once to obtain a fixed embedding, and a logistic-regression head was trained on these embeddings under the same 10-fold stratified cross-validation used for the trained models. Image-only, text-only, and combined image-plus-text probes were fitted wherever the model exposed the corresponding encoder.

### Performance metrics

Trained vision and language models were assessed by 10-fold stratified cross-validation with a shared fold seed. For each model we report accuracy (ACC), sensitivity (SEN), specificity (SPE), F1, and the area under the receiver operating characteristic curve (AUC). Per-fold values were aggregated as mean ± standard deviation with *t*-distribution-based 95% confidence intervals (CI), and a pooled confusion matrix, reporting true negatives (TN), false positives (FP), false negatives (FN), and true positives (TP), was computed.^25^ The VLMs were evaluated under the same protocol using frozen-encoder linear probes. Abnormal was treated as the positive class throughout.

### Parameters specification and implementation

Experiments were implemented in PyTorch and run on the Apocrita high-performance computing cluster (Queen Mary University of London) using NVIDIA Tesla V100 and A100/H100 GPUs.

The eight vision architectures were fine-tuned from ImageNet-pretrained weights for up to 50 epochs with the Adam optimiser (learning rate 1×10⁻⁴) and a batch size of 16 (reduced to 8 for the two neural-architecture-search networks); a fixed random seed (42) was used throughout, with the same 10-fold stratified cross-validation and fold assignment applied to every model.

The seven language classifiers used the same fixed 10-fold assignment and seed (42). The five transformer models (BERT-base, RoBERTa-base, DistilBERT, BioBERT, Bio_ClinicalBERT) were fine-tuned for 5 epochs with the AdamW optimiser (learning rate 2×10⁻⁵), a batch size of 16, and a maximum sequence length of 256 tokens. BiLSTM and TextCNN, whose embeddings are learned from scratch, were trained for 20 epochs with the Adam optimiser (learning rate 1×10⁻³), a batch size of 16, a 128-dimensional embedding layer, a hidden size of 128, and the same 256-token maximum length via a regular-expression word tokeniser. No early-stopping criterion was applied to any language model; all seven trained for a fixed number of epochs.

For the VLM linear-probe arm, image and text embeddings were extracted once from each frozen encoder (CLIP ViT-B/16, BiomedCLIP, Qwen2·5-VL, MedGemma-4B, LLaVA-Med v1·5), combining the pooled vision-tower output with a mean-pooled hidden-state text embedding; because the encoders are frozen, embeddings do not depend on the label condition and were reused for both the pre and post runs. Embeddings were standardised (zero mean, unit variance) within each training fold, and a logistic-regression probe (scikit-learn; L2 penalty, C=1·0, lbfgs solver, no class weighting, maximum 5000 iterations) was fitted on the standardised training folds and evaluated on the held-out fold, using the same fixed 10-fold assignment and seed (42) as the vision and language arms.

Gradient-weighted class activation mapping (Grad-CAM)²⁶ was used to visualise the image regions driving the convolutional models’ predictions: the pytorch_grad_cam implementation was used for the five standard CNN architectures, and a custom forward/backward-hook implementation with in-place activations disabled was used for the two neural-architecture-search networks (NASNet-Large, PNASNet-5-Large), whose in-place layers are otherwise incompatible with backward-hook-based Grad-CAM; the target layer for every architecture was its last convolutional block producing a spatial feature map.

## Results

### Image-only and language model classification

Across the eight imaging architectures, AUC ranged from 0·799 to 0·880 (Table 3). PNASNet-5-Large was the strongest (AUC 0·880, 95% CI 0·850–0·910), followed closely by DenseNet-121 and EfficientNet-B0 (AUC 0·863 each); EfficientNet-B0 achieved the highest accuracy (0·800) with the most balanced sensitivity/specificity profile (0·787/0·813). The convolutional and NAS networks clustered tightly (AUC 0·844–0·880), indicating that architecture choice within the CNN family had a modest effect at this sample size. AlexNet was the weakest convolutional model (accuracy 0·740; sensitivity 0·680). Grad-CAM activation maps for representative examples from each network are shown in Figure 3.

**Figure 3.**
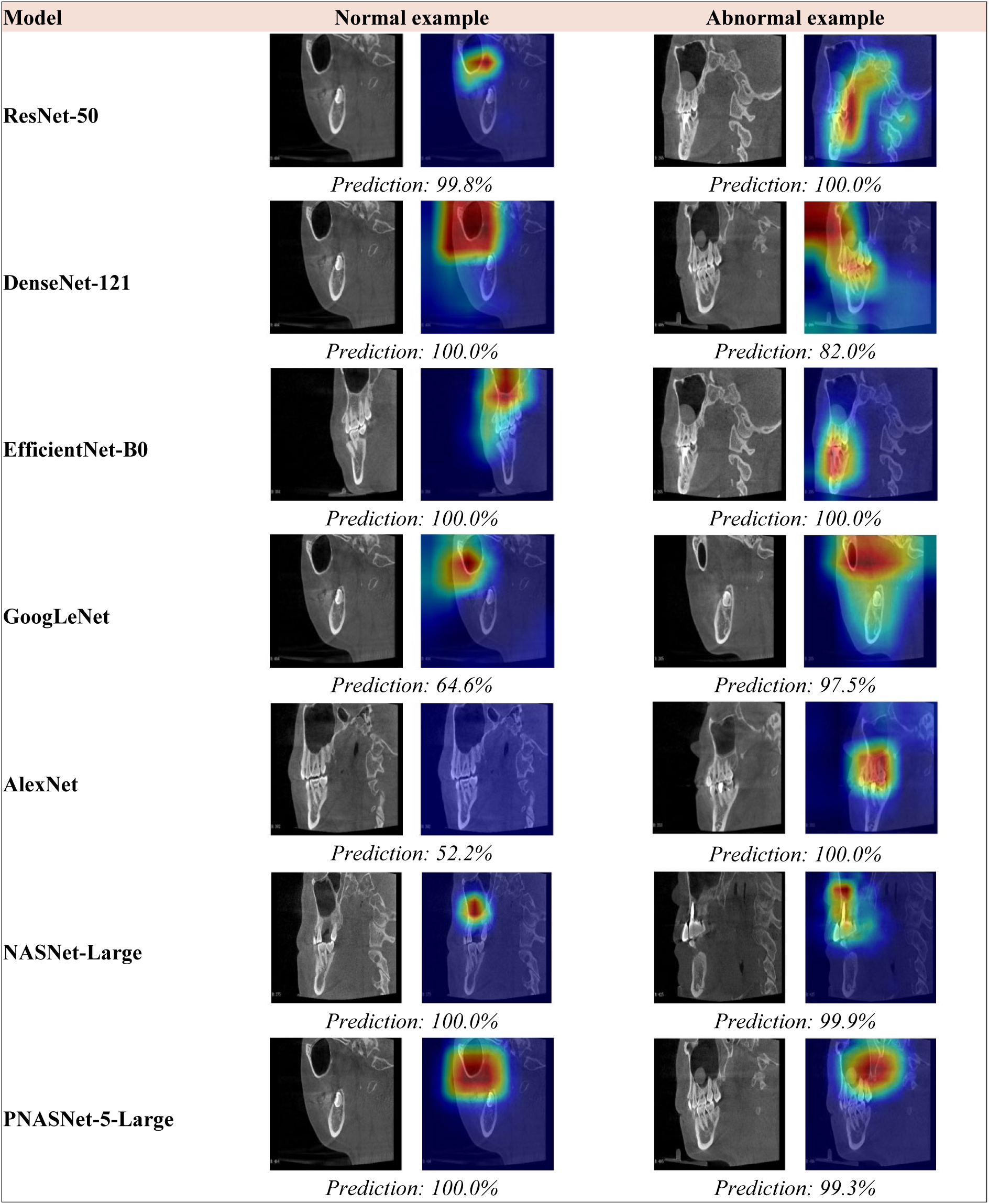
Grad-CAM activation maps for representative convolutional and NAS models, showing one correctly classified normal and one correctly classified abnormal CBCT example per model. Heatmap intensities indicate the image regions most influential to each model’s prediction.

**Table 3.** Combined classification performance across the vision, language, and VLM families. For the trained vision and language models, each cell reports the mean ± standard deviation across the 10 stratified cross-validation folds, with the t-distribution-based 95% confidence interval in parentheses.

| Model | ACC | SEN | SPE | F1 | AUC |
| --- | --- | --- | --- | --- | --- |
| **Vision models** |  |  |  |  |  |
| ResNet-50 | 0.760 ± 0.072<br>(0.709–0.811) | 0.753 ± 0.104<br>(0.679–0.828) | 0.767 ± 0.194<br>(0.628–0.906) | 0.760 ± 0.057<br>(0.719–0.800) | 0.844 ± 0.053<br>(0.806–0.882) |
| DenseNet-121 | 0.760 ± 0.083<br>(0.701–0.819) | 0.720 ± 0.203<br>(0.575–0.865) | 0.800 ± 0.126<br>(0.710–0.890) | 0.734 ± 0.155<br>(0.623–0.844) | 0.863 ± 0.051<br>(0.826–0.899) |
| EfficientNet-B0 | 0.800 ± 0.059<br>(0.758–0.842) | 0.787 ± 0.098<br>(0.716–0.857) | 0.813 ± 0.117<br>(0.730–0.897) | 0.796 ± 0.060<br>(0.753–0.839) | 0.863 ± 0.047<br>(0.830–0.896) |
| GoogLeNet | 0.787 ± 0.050<br>(0.751–0.823) | 0.727 ± 0.097<br>(0.658–0.796) | 0.847 ± 0.109<br>(0.769–0.925) | 0.771 ± 0.058<br>(0.730–0.813) | 0.862 ± 0.060<br>(0.819–0.905) |
| AlexNet | 0.740 ± 0.098<br>(0.670–0.810) | 0.680 ± 0.174<br>(0.555–0.805) | 0.800 ± 0.154<br>(0.690–0.910) | 0.715 ± 0.121<br>(0.628–0.802) | 0.852 ± 0.081<br>(0.794–0.909) |
| NASNet-Large | 0.780 ± 0.028<br>(0.760–0.800) | 0.707 ± 0.105<br>(0.631–0.782) | 0.853 ± 0.088<br>(0.790–0.916) | 0.759 ± 0.048<br>(0.725–0.794) | 0.857 ± 0.031<br>(0.835–0.879) |
| PNASNet-5-Large | 0.790 ± 0.070<br>(0.740–0.840) | 0.753 ± 0.134<br>(0.658–0.849) | 0.827 ± 0.148<br>(0.721–0.933) | 0.779 ± 0.082<br>(0.721–0.838) | 0.880 ± 0.041<br>(0.850–0.910) |
| ViT-B/16 | 0.700 ± 0.067<br>(0.652–0.748) | 0.540 ± 0.244<br>(0.365–0.715) | 0.860 ± 0.149<br>(0.753–0.967) | 0.610 ± 0.179<br>(0.482–0.739) | 0.799 ± 0.056<br>(0.759–0.839) |
| **Language models** |  |  |  |  |  |
| BiLSTM | 0.977 ± 0.039<br>(0.949–1.004) | 0.987 ± 0.028<br>(0.967–1.007) | 0.967 ± 0.065<br>(0.920–1.013) | 0.978 ± 0.037<br>(0.951–1.004) | 0.998 ± 0.004<br>(0.995–1.001) |
| TextCNN | 0.987 ± 0.028<br>(0.967–1.007) | 0.987 ± 0.028<br>(0.967–1.007) | 0.987 ± 0.028<br>(0.967–1.007) | 0.987 ± 0.028<br>(0.967–1.007) | 0.999 ± 0.003<br>(0.997–1.001) |
| BERT-base | 0.993 ± 0.014<br>(0.983–1.003) | 0.987 ± 0.028<br>(0.967–1.007) | 1.000 ± 0.000<br>(1.000–1.000) | 0.993 ± 0.015<br>(0.983–1.004) | 0.998 ± 0.006<br>(0.994–1.002) |
| RoBERTa-base | 0.983 ± 0.024<br>(0.967–1.000) | 0.967 ± 0.047<br>(0.933–1.000) | 1.000 ± 0.000<br>(1.000–1.000) | 0.983 ± 0.025<br>(0.965–1.000) | 0.992 ± 0.019<br>(0.978–1.005) |
| DistilBERT | 0.993 ± 0.014<br>(0.983–1.003) | 0.987 ± 0.028<br>(0.967–1.007) | 1.000 ± 0.000<br>(1.000–1.000) | 0.993 ± 0.015<br>(0.983–1.004) | 0.999 ± 0.002<br>(0.998–1.000) |
| BioBERT | 0.993 ± 0.014<br>(0.983–1.003) | 0.987 ± 0.028<br>(0.967–1.007) | 1.000 ± 0.000<br>(1.000–1.000) | 0.993 ± 0.015<br>(0.983–1.004) | 0.999 ± 0.002<br>(0.998–1.000) |
| Bio_ClinicalBERT | 0.990 ± 0.022<br>(0.974–1.006) | 0.987 ± 0.028<br>(0.967–1.007) | 0.993 ± 0.021<br>(0.978–1.008) | 0.990 ± 0.023<br>(0.974–1.006) | 1.000 ± 0.001<br>(0.999–1.001) |
| **VLMs** |  |  |  |  |  |
| Qwen2.5-VL (image only) | 0.690 ± 0.069<br>(0.641–0.739) | 0.700 ± 0.096<br>(0.632–0.768) | 0.680 ± 0.129<br>(0.588–0.772) | 0.692 ± 0.063<br>(0.647–0.738) | 0.750 ± 0.099<br>(0.679–0.821) |
| Qwen2.5-VL (text only) | 0.540 ± 0.031<br>(0.518–0.562) | 0.080 ± 0.061<br>(0.036–0.124) | 1.000 ± 0.000<br>(1.000–1.000) | 0.143 ± 0.103<br>(0.069–0.217) | 0.541 ± 0.036<br>(0.516–0.567) |
| Qwen2.5-VL (image+text) | 0.700 ± 0.069<br>(0.651–0.749) | 0.707 ± 0.118<br>(0.622–0.791) | 0.693 ± 0.100<br>(0.622–0.765) | 0.699 ± 0.077<br>(0.644–0.754) | 0.777 ± 0.097<br>(0.708–0.846) |
| MedGemma-4B (image only) | 0.803 ± 0.066<br>(0.756–0.850) | 0.800 ± 0.089<br>(0.736–0.864) | 0.807 ± 0.102<br>(0.734–0.879) | 0.802 ± 0.065<br>(0.756–0.849) | 0.874 ± 0.049<br>(0.839–0.909) |
| MedGemma-4B (text only) | 0.540 ± 0.031<br>(0.518–0.562) | 0.080 ± 0.061<br>(0.036–0.124) | 1.000 ± 0.000<br>(1.000–1.000) | 0.143 ± 0.103<br>(0.069–0.217) | 0.552 ± 0.043<br>(0.521–0.583) |
| MedGemma-4B (image+text) | 0.810 ± 0.061<br>(0.766–0.854) | 0.807 ± 0.106<br>(0.731–0.883) | 0.813 ± 0.103<br>(0.740–0.887) | 0.808 ± 0.064<br>(0.762–0.853) | 0.884 ± 0.052<br>(0.847–0.921) |
| LLaVA-Med v1.5 (image only) | 0.713 ± 0.048<br>(0.679–0.747) | 0.727 ± 0.102<br>(0.654–0.799) | 0.700 ± 0.110<br>(0.621–0.779) | 0.715 ± 0.053<br>(0.677–0.753) | 0.796 ± 0.069<br>(0.747–0.846) |
| LLaVA-Med v1.5 (text only) | 0.987 ± 0.017<br>(0.974–0.999) | 0.987 ± 0.028<br>(0.967–1.000) | 0.987 ± 0.028<br>(0.967–1.000) | 0.987 ± 0.017<br>(0.974–0.999) | 1.000 ± 0.000<br>(1.000–1.000) |
| LLaVA-Med v1.5 (image+text) | $0.987 \pm 0.017$<br>(0.974–0.999) | $0.987 \pm 0.028$<br>(0.967–1.000) | $0.987 \pm 0.028$<br>(0.967–1.000) | $0.987 \pm 0.017$<br>(0.974–0.999) | $1.000 \pm 0.000$<br>(1.000–1.000) |
| BiomedCLIP (image only) | $0.587 \pm 0.076$<br>(0.533–0.641) | $0.607 \pm 0.135$<br>(0.510–0.703) | $0.567 \pm 0.134$<br>(0.471–0.663) | $0.590 \pm 0.093$<br>(0.524–0.656) | $0.628 \pm 0.083$<br>(0.569–0.688) |
| BiomedCLIP (text only) | $0.983 \pm 0.018$<br>(0.971–0.996) | $0.980 \pm 0.032$<br>(0.957–1.000) | $0.987 \pm 0.028$<br>(0.967–1.000) | $0.983 \pm 0.018$<br>(0.971–0.996) | $1.000 \pm 0.000$<br>(1.000–1.000) |
| BiomedCLIP (image+text) | $0.993 \pm 0.014$<br>(0.983–1.000) | $1.000 \pm 0.000$<br>(1.000–1.000) | $0.987 \pm 0.028$<br>(0.967–1.000) | $0.994 \pm 0.014$<br>(0.984–1.000) | $1.000 \pm 0.000$<br>(1.000–1.000) |
| CLIP ViT-B/16 (image only) | $0.640 \pm 0.072$<br>(0.589–0.691) | $0.653 \pm 0.093$<br>(0.587–0.720) | $0.627 \pm 0.130$<br>(0.533–0.720) | $0.645 \pm 0.066$<br>(0.598–0.691) | $0.685 \pm 0.067$<br>(0.638–0.733) |
| CLIP ViT-B/16 (text only) | $0.983 \pm 0.018$<br>(0.971–0.996) | $0.987 \pm 0.028$<br>(0.967–1.000) | $0.980 \pm 0.032$<br>(0.957–1.000) | $0.983 \pm 0.018$<br>(0.971–0.996) | $1.000 \pm 0.001$<br>(0.999–1.000) |
| CLIP ViT-B/16 (image+text) | $0.983 \pm 0.018$<br>(0.971–0.996) | $0.987 \pm 0.028$<br>(0.967–1.000) | $0.980 \pm 0.032$<br>(0.957–1.000) | $0.983 \pm 0.018$<br>(0.971–0.996) | $1.000 \pm 0.000$<br>(1.000–1.000) |

ViT-B/16 underperformed all other CNNs (ACC 0·700; AUC 0·799). Its errors were strongly asymmetric: specificity was high (0·860) but sensitivity was low (0·540), with 69 abnormal sinuses misclassified as normal (pooled confusion matrix 129/21/69/81 for TN/FP/FN/TP). This pattern, high specificity, poor sensitivity, is the expected signature of a data-hungry architecture trained on only a few hundred images.

In striking contrast, all seven language models reached near-ceiling performance on the findings text (accuracy 0·977–0·993; AUC 0·992–1·000; Table 3). Bio_ClinicalBERT attained a perfect AUC (1·000), and BERT-base, DistilBERT, and BioBERT each reached accuracy 0·993 with only two misclassifications out of 300. This uniformity reflects the diagnostic of the findings text: normal cases are described as a list of absences while abnormal cases name the pathology. Because seven architecturally diverse models all converge near 100%, the discriminative information is carried by the findings text itself rather than by any model’s reasoning, and these scores are not directly comparable with the imaging AUCs.

### Generative note drafting

All three generative VLMs produced a structured findings/impression note for every case (representative outputs in Figure 2). The models differed markedly in their level of detail and reliability. Qwen2·5-VL generated the longest notes (mean 161 words vs 78 for MedGemma-4B and 88 for LLaVA-Med v1·5), and 66% ended without sentence-final punctuation, consistent with reaching the generation-length limit. The clinically important difference was reliability: among abnormal cases whose input did not specify a side, Qwen2·5-VL introduced an unsupported laterality in 17 of 150 cases (11%), MedGemma-4B in 4 of 150 (3%), and LLaVA-Med v1·5 in none. MedGemma-4B reproduced an unfilled clinical template with placeholder fields in 200 of 300 notes (67%).

LLaVA-Med v1·5 was the most conservative: short, complete, and free of invented detail, making it the safest default for findings-conditioned note drafting. Qwen2·5-VL is the most verbose but occasionally asserts findings the input did not contain, which is unsafe for unsupervised documentation. MedGemma-4B is concise but defaults to an unfilled template requiring a clean-up step before clinical use.

### VLMs under linear probing

Under frozen-encoder linear probing (same 10-fold protocol as Table 3), the three generative VLMs showed two distinct patterns. All carried genuine image-only diagnostic signal: MedGemma-4B was strongest (image-only AUC 0·874, 95% CI 0·839–0·909; accuracy 0·803), approaching the trained CNNs; LLaVA-Med v1·5 (AUC 0·796) and Qwen2·5-VL (0·750) were both well above chance. The conditions diverged on text: for Qwen2·5-VL and MedGemma-4B the text-only probe collapsed near chance (AUC 0·541 and 0·552), so diagnostic signal is carried almost entirely by the image. LLaVA-Med v1·5 behaved oppositely, its text embeddings showed exceptionally high separability (text-only AUC 1·000, accuracy 0·987), mirroring the language-model arm.

The two contrastive models showed the weakest image-only probes (CLIP ViT-B/16 AUC 0·685, BiomedCLIP 0·628) but exceptionally high-accuracy text-only probes (both AUC ≥0·99), grouping them with the language models and LLaVA-Med on the findings-text side of the comparison. Figure 4 summarises AUC by model family.

**Figure 4.**
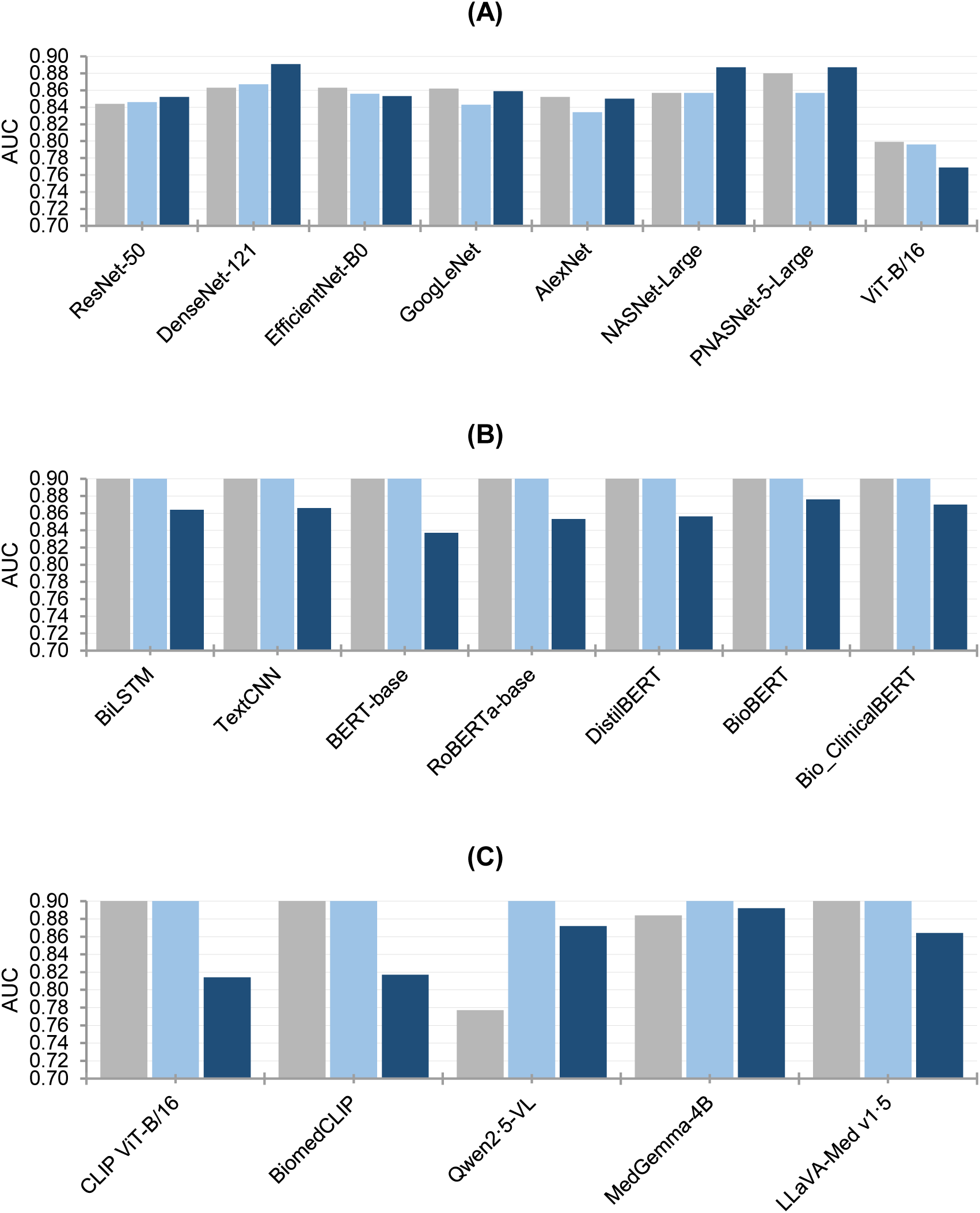
Classification performance (AUC) across the vision (A), language (B), and VLM arms (C), grouped by model family. For each model, three bars show mean AUC using ChatGPT’s own labels alone before radiologist review, on the full 300-case dataset (grey); the provisional ChatGPT-only labels after excluding the three cases with insufficient source material, n=297 (light blue); and the radiologist-confirmed labels, in which 62 of the 297 cases were reclassified relative to the ChatGPT-only label (dark blue).

### Pre- versus post-radiologist label sensitivity analysis

To quantify the contribution of the radiologist’s label corrections, all seven language models were retrained under an identical 10-fold protocol on two label conditions drawn from the same fixed case-to-fold assignment: the provisional, ChatGPT-only labels (“pre”) and the same cases’ radiologist-confirmed labels after their review (“post”). Every model showed a consistent, substantial drop in AUC from the near-ceiling pre-correction values, indicating that a meaningful fraction of the apparent language-model performance in the main analysis reflected agreement with the provisional ChatGPT output rather than with the true reference standard (Table 4).

**Table 4.** Language-model performance under identical 10-fold stratified cross-validation and fixed fold assignment, comparing the provisional ChatGPT-only labels (“pre”) against the radiologist-confirmed labels (“post”) for the same seven models. Values are mean ± standard deviation across folds.

| Model | ACC | SEN | SPE | F1 | AUC |
| --- | --- | --- | --- | --- | --- |
| *Pre—provisional ChatGPT-only labels* |  |  |  |  |  |
| BiLSTM | 0.984 ± 0.032 | 0.973 ± 0.046 | 0.993 ± 0.021 | 0.983 ± 0.032 | 0.999 ± 0.003 |
| TextCNN | 0.987 ± 0.017 | 0.986 ± 0.03 | 0.986 ± 0.029 | 0.986 ± 0.018 | 0.999 ± 0.003 |
| BERT-base | 0.99 ± 0.016 | 0.986 ± 0.03 | 0.993 ± 0.023 | 0.99 ± 0.017 | 0.999 ± 0.004 |
| RoBERTa-base | 0.99 ± 0.016 | 0.986 ± 0.03 | 0.993 ± 0.023 | 0.99 ± 0.017 | 1.0 ± 0.001 |
| DistilBERT | 0.984 ± 0.032 | 0.973 ± 0.046 | 0.993 ± 0.021 | 0.983 ± 0.032 | 0.999 ± 0.004 |
| BioBERT | 0.99 ± 0.016 | 0.986 ± 0.03 | 0.993 ± 0.023 | 0.99 ± 0.017 | 1.0 ± 0.0 |
| Bio_ClinicalBERT | 0.99 ± 0.016 | 0.98 ± 0.033 | 1.0 ± 0.0 | 0.989 ± 0.017 | 1.0 ± 0.0 |
| *Post—radiologist-confirmed labels* |  |  |  |  |  |
| BiLSTM | 0.804 ± 0.059 | 0.741 ± 0.112 | 0.869 ± 0.101 | 0.791 ± 0.077 | 0.864 ± 0.063 |
| TextCNN | 0.771 ± 0.077 | 0.809 ± 0.108 | 0.732 ± 0.158 | 0.782 ± 0.073 | 0.866 ± 0.088 |
| BERT-base | 0.812 ± 0.065 | 0.741 ± 0.098 | 0.883 ± 0.093 | 0.798 ± 0.076 | 0.837 ± 0.094 |
| RoBERTa-base | 0.795 ± 0.041 | 0.676 ± 0.12 | 0.919 ± 0.108 | 0.766 ± 0.064 | 0.853 ± 0.09 |
| DistilBERT | 0.792 ± 0.079 | 0.715 ± 0.078 | 0.871 ± 0.106 | 0.778 ± 0.084 | 0.856 ± 0.097 |
| BioBERT | 0.818 ± 0.053 | 0.741 ± 0.107 | 0.896 ± 0.077 | 0.802 ± 0.075 | 0.876 ± 0.073 |
| Bio_ClinicalBERT | 0.805 ± 0.076 | 0.695 ± 0.181 | 0.918 ± 0.054 | 0.768 ± 0.155 | 0.87 ± 0.102 |

The same pre/post comparison was repeated for all eight vision architectures, retrained under identical parameters on the two label conditions with the same fixed fold assignment. In sharp contrast to the language arm, vision AUC changed only marginally between conditions (Table 5): six of the eight models showed a small AUC increase after label correction (+0·006 to +0·030), consistent with training and evaluating against the true rather than provisional diagnosis; EfficientNet-B0 was essentially unchanged (−0·003), and ViT-B/16 showed a small decrease (−0·027), in keeping with its already-noted instability at this sample size. No vision model showed anything resembling the language arm’s uniform, large performance drop. This asymmetry indicates that the trained vision models derive their discriminative signal predominantly from the images themselves rather than from label-provenance artefacts, whereas the language arm’s near-ceiling performance is substantially attributable to the specific label set used for training and evaluation.

**Table 5.**
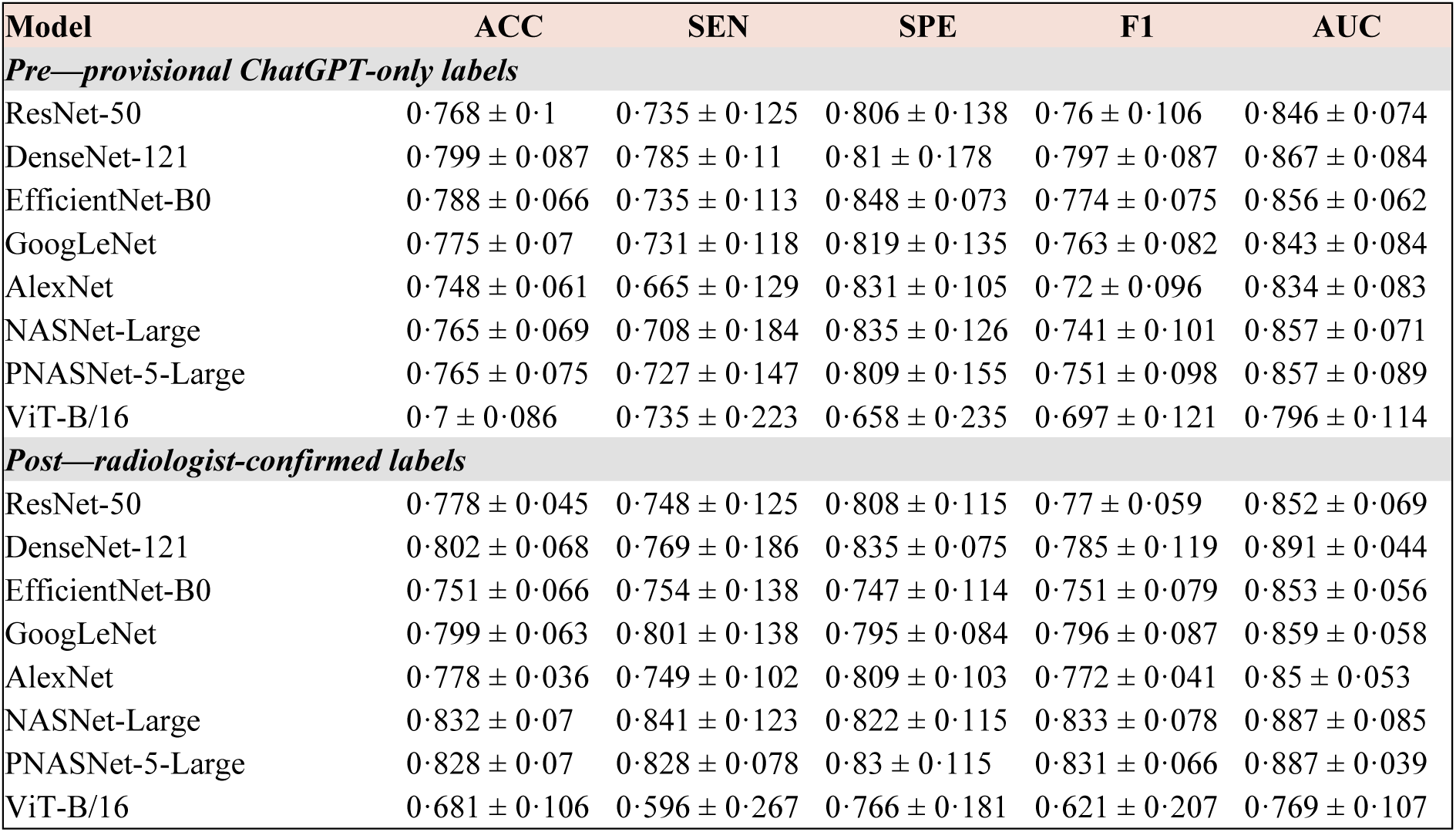
Vision-model performance under identical 10-fold stratified cross-validation and fixed fold assignment, comparing the provisional ChatGPT-only labels (“pre”) against the radiologist-confirmed labels (“post”) for the same eight architectures. Values are mean ± standard deviation across folds.

| Model | ACC | SEN | SPE | F1 | AUC |
| --- | --- | --- | --- | --- | --- |
| *Pre—provisional ChatGPT-only labels* |  |  |  |  |  |
| ResNet-50 | 0.768 ± 0.1 | 0.735 ± 0.125 | 0.806 ± 0.138 | 0.76 ± 0.106 | 0.846 ± 0.074 |
| DenseNet-121 | 0.799 ± 0.087 | 0.785 ± 0.11 | 0.81 ± 0.178 | 0.797 ± 0.087 | 0.867 ± 0.084 |
| EfficientNet-B0 | 0.788 ± 0.066 | 0.735 ± 0.113 | 0.848 ± 0.073 | 0.774 ± 0.075 | 0.856 ± 0.062 |
| GoogLeNet | 0.775 ± 0.07 | 0.731 ± 0.118 | 0.819 ± 0.135 | 0.763 ± 0.082 | 0.843 ± 0.084 |
| AlexNet | 0.748 ± 0.061 | 0.665 ± 0.129 | 0.831 ± 0.105 | 0.72 ± 0.096 | 0.834 ± 0.083 |
| NASNet-Large | 0.765 ± 0.069 | 0.708 ± 0.184 | 0.835 ± 0.126 | 0.741 ± 0.101 | 0.857 ± 0.071 |
| PNASNet-5-Large | 0.765 ± 0.075 | 0.727 ± 0.147 | 0.809 ± 0.155 | 0.751 ± 0.098 | 0.857 ± 0.089 |
| ViT-B/16 | 0.7 ± 0.086 | 0.735 ± 0.223 | 0.658 ± 0.235 | 0.697 ± 0.121 | 0.796 ± 0.114 |
| *Post—radiologist-confirmed labels* |  |  |  |  |  |
| ResNet-50 | 0.778 ± 0.045 | 0.748 ± 0.125 | 0.808 ± 0.115 | 0.77 ± 0.059 | 0.852 ± 0.069 |
| DenseNet-121 | 0.802 ± 0.068 | 0.769 ± 0.186 | 0.835 ± 0.075 | 0.785 ± 0.119 | 0.891 ± 0.044 |
| EfficientNet-B0 | 0.751 ± 0.066 | 0.754 ± 0.138 | 0.747 ± 0.114 | 0.751 ± 0.079 | 0.853 ± 0.056 |
| GoogLeNet | 0.799 ± 0.063 | 0.801 ± 0.138 | 0.795 ± 0.084 | 0.796 ± 0.087 | 0.859 ± 0.058 |
| AlexNet | 0.778 ± 0.036 | 0.749 ± 0.102 | 0.809 ± 0.103 | 0.772 ± 0.041 | 0.85 ± 0.053 |
| NASNet-Large | 0.832 ± 0.07 | 0.841 ± 0.123 | 0.822 ± 0.115 | 0.833 ± 0.078 | 0.887 ± 0.085 |
| PNASNet-5-Large | 0.828 ± 0.07 | 0.828 ± 0.078 | 0.83 ± 0.115 | 0.831 ± 0.066 | 0.887 ± 0.039 |
| ViT-B/16 | 0.681 ± 0.106 | 0.596 ± 0.267 | 0.766 ± 0.181 | 0.621 ± 0.207 | 0.769 ± 0.107 |

The same pre/post comparison was extended to the five VLMs under frozen-encoder linear probing, across all three input conditions (image-only, text-only, image-plus-text; Table 6). The pattern mirrored the two trained arms exactly. Text-only and image-plus-text probes, exceptionally high on the provisional labels for every model (AUC ≥0·99), fell sharply after label correction (e.g. CLIP ViT-B/16 text-only AUC 0·999 to 0·841; MedGemma-4B image-plus-text 1·000 to 0·892), matching the collapse already seen in the standalone language arm. Image-only probes, by contrast, changed only modestly (e.g. MedGemma-4B 0·875 to 0·835; Qwen2·5-VL 0·741 to 0·723), consistent with the vision arm’s relative robustness. This three-way replication—vision, language, and every VLM’s text-derived probe sensitive— reinforces that apparent multimodal competence in this benchmark tracks the specific label set used for the findings text, not independent image understanding.

**Table 6.** VLM linear-probe performance for each of the five VLMs under the image-only, text-only, and image-plus-text probe conditions, using the same 10-fold stratified cross-validation and fixed fold assignment throughout, comparing the provisional ChatGPT-only labels (“pre”) against the radiologist-confirmed labels (“post”). Values are mean ± standard deviation across folds.

| Model (probe condition) | ACC | SEN | SPE | F1 | AUC |
| --- | --- | --- | --- | --- | --- |
| *Pre—provisional ChatGPT-only labels* |  |  |  |  |  |
| CLIP-ViT-B16 (Image only) | 0.627 ± 0.068 | 0.624 ± 0.109 | 0.620 ± 0.106 | 0.622 ± 0.091 | 0.692 ± 0.082 |
| CLIP-ViT-B16 (Text only) | 0.990 ± 0.016 | 0.986 ± 0.030 | 0.993 ± 0.021 | 0.990 ± 0.017 | 0.999 ± 0.003 |
| CLIP-ViT-B16 (Image + text) | 0.990 ± 0.016 | 0.986 ± 0.030 | 0.993 ± 0.021 | 0.990 ± 0.017 | 1.000 ± 0.000 |
| BiomedCLIP (Image only) | 0.573 ± 0.086 | 0.584 ± 0.127 | 0.562 ± 0.163 | 0.575 ± 0.092 | 0.639 ± 0.089 |
| BiomedCLIP (Text only) | 0.990 ± 0.016 | 0.986 ± 0.030 | 0.993 ± 0.021 | 0.990 ± 0.017 | 0.999 ± 0.003 |
| BiomedCLIP (Image + text) | 0.997 ± 0.010 | 1.000 ± 0.000 | 0.993 ± 0.021 | 0.997 ± 0.010 | 1.000 ± 0.000 |
| Qwen2.5-VL (Image only) | 0.698 ± 0.085 | 0.696 ± 0.115 | 0.695 ± 0.090 | 0.695 ± 0.098 | 0.741 ± 0.081 |
| Qwen2.5-VL (Text only) | 0.990 ± 0.016 | 0.986 ± 0.030 | 0.993 ± 0.021 | 0.990 ± 0.017 | 0.998 ± 0.007 |
| Qwen2.5-VL (Image + text) | 0.987 ± 0.024 | 0.979 ± 0.048 | 0.993 ± 0.021 | 0.986 ± 0.026 | 1.000 ± 0.001 |
| MedGemma-4B (Image only) | 0.782 ± 0.083 | 0.786 ± 0.096 | 0.778 ± 0.132 | 0.783 ± 0.085 | 0.875 ± 0.049 |
| MedGemma-4B (Text only) | 0.990 ± 0.016 | 0.986 ± 0.030 | 0.993 ± 0.021 | 0.990 ± 0.017 | 0.998 ± 0.005 |
| MedGemma-4B (Image + text) | 0.990 ± 0.016 | 0.986 ± 0.030 | 0.993 ± 0.021 | 0.990 ± 0.017 | 1.000 ± 0.000 |
| LLaVA-Med v1.5 (Image only) | 0.701 ± 0.044 | 0.701 ± 0.136 | 0.705 ± 0.121 | 0.696 ± 0.060 | 0.803 ± 0.035 |
| LLaVA-Med v1.5 (Text only) | 0.990 ± 0.016 | 0.986 ± 0.030 | 0.993 ± 0.021 | 0.990 ± 0.017 | 0.998 ± 0.005 |
| LLaVA-Med v1.5 (Image + text) | 0.987 ± 0.017 | 0.986 ± 0.030 | 0.986 ± 0.029 | 0.987 ± 0.018 | 1.000 ± 0.000 |
| *Post—radiologist-confirmed labels* |  |  |  |  |  |
| CLIP-ViT-B16 (Image only) | 0.615 ± 0.070 | 0.596 ± 0.080 | 0.636 ± 0.088 | 0.611 ± 0.072 | 0.649 ± 0.072 |
| CLIP-ViT-B16 (Text only) | 0.781 ± 0.059 | 0.722 ± 0.137 | 0.843 ± 0.094 | 0.764 ± 0.089 | 0.841 ± 0.086 |
| CLIP-ViT-B16 (Image + text) | 0.744 ± 0.096 | 0.735 ± 0.126 | 0.754 ± 0.147 | 0.744 ± 0.103 | 0.814 ± 0.089 |
| BiomedCLIP (Image only) | 0.566 ± 0.101 | 0.603 ± 0.144 | 0.528 ± 0.131 | 0.581 ± 0.114 | 0.612 ± 0.122 |
| BiomedCLIP (Text only) | 0.774 ± 0.069 | 0.715 ± 0.131 | 0.837 ± 0.102 | 0.758 ± 0.095 | 0.831 ± 0.078 |
| BiomedCLIP (Image + text) | 0.748 ± 0.088 | 0.728 ± 0.135 | 0.769 ± 0.127 | 0.742 ± 0.098 | 0.817 ± 0.090 |
| Qwen2.5-VL (Image only) | 0.668 ± 0.063 | 0.682 ± 0.115 | 0.652 ± 0.108 | 0.673 ± 0.074 | 0.723 ± 0.080 |
| Qwen2.5-VL (Text only) | 0.781 ± 0.057 | 0.755 ± 0.114 | 0.809 ± 0.089 | 0.775 ± 0.076 | 0.856 ± 0.073 |
| Qwen2.5-VL (Image + text) | 0.808 ± 0.074 | 0.801 ± 0.110 | 0.817 ± 0.141 | 0.808 ± 0.074 | 0.872 ± 0.050 |
| MedGemma-4B (Image only) | 0.768 ± 0.095 | 0.801 ± 0.055 | 0.736 ± 0.183 | 0.782 ± 0.074 | 0.835 ± 0.078 |
| MedGemma-4B (Text only) | 0.761 ± 0.062 | 0.728 ± 0.116 | 0.795 ± 0.119 | 0.753 ± 0.074 | 0.841 ± 0.070 |
| MedGemma-4B (Image + text) | 0.825 ± 0.075 | 0.828 ± 0.110 | 0.822 ± 0.133 | 0.827 ± 0.076 | 0.892 ± 0.043 |
| LLaVA-Med v1.5 (Image only) | 0.731 ± 0.077 | 0.761 ± 0.091 | 0.701 ± 0.130 | 0.742 ± 0.072 | 0.771 ± 0.087 |
| LLaVA-Med v1.5 (Text only) | 0.767 ± 0.069 | 0.735 ± 0.110 | 0.801 ± 0.108 | 0.760 ± 0.082 | 0.840 ± 0.077 |
| LLaVA-Med v1.5 (Image + text) | 0.798 ± 0.053 | 0.787 ± 0.099 | 0.809 ± 0.111 | 0.797 ± 0.062 | 0.864 ± 0.062 |

## Discussion

Three findings stand out. The CNNs learned the task well, with PNASNet-5-Large reaching AUC 0·880 and EfficientNet-B0 reaching 0·800 accuracy. The radiologist-verified findings text drove every language model to exceptionally high scores. VLMs probed with a linear head on frozen features recovered only modest image-only signal while classifying the same findings text almost perfectly. Together, these results convey a single coherent message: supervised imaging works, the written findings are themselves highly predictive, and image-only multimodal signal remains weak for this appearance.

The pre/post comparison clarifies the trade-offs between the two label sources rather than establishing as unconditionally superior. ChatGPT-only labelling offered clear practical advantages: it was fast, inexpensive, and scalable, and produced consistently formatted findings text without requiring specialist time. However, this convenience came at the cost of construct validity for the language and VLM-text arms. Every language model reached exceptionally high AUC on ChatGPT-only labels (0·99–1·00; Table 4), yet these scores fell substantially once evaluated against the radiologist-confirmed reference standard (e.g. BERT-base 0·999 to 0·837, RoBERTa-base 1·000 to 0·853), indicating that much of the apparent accuracy reflected models learning to reproduce ChatGPT’s own labelling patterns rather than genuine diagnostic signal. Radiologist confirmation is costlier and slower to obtain but functions as the true clinical ground truth and is therefore the only condition under which language-arm performance can be considered clinically meaningful. The vision arm told a different story: most CNN architectures were stable or modestly improved under radiologist-confirmed labels (e.g. DenseNet-121 0·867 to 0·891, PNASNet-5-Large 0·857 to 0·887; Table 5), consistent with these models learning from image content that the label source does not alter. The same asymmetry recurred within the VLM-probe arm, where image-only performance moved little (e.g. MedGemma-4B 0·875 to 0·835) while text-derived probes collapsed alongside the language arm (e.g. MedGemma-4B image-plus-text 1·000 to 0·892; Table 6).

The underperformance of ViT-B/16 relative to every CNN is consistent with transformers’ well-documented dependence on large training corpora and confirms that this dependence holds even at the modest dataset scale typical of dental CBCT research. ViTs lack the inductive biases of convolution and must learn spatial structure from data; with only a few hundred training images they cannot do so reliably, manifesting here as poor sensitivity (0·540) and many missed abnormal sinuses. For dental-imaging datasets of realistic size, convolutional and NAS networks remain the more dependable default, and reporting a transformer baseline alongside them guards against overstating its readiness.

Grad-CAM maps (Figure 3) support this reliability qualitatively for most networks: activations concentrated on the maxillary sinus itself, over the air-filled cavity in normal cases and over the opacified or lesional region in abnormal cases, rather than on surrounding teeth or cortical bone. Localisation was tightest for NASNet-Large and PNASNet-5-Large. AlexNet was an exception: despite predicting the abnormal example correctly, its activation concentrated on the dentition rather than the sinus cavity, suggesting its correct prediction may have relied partly on a non-sinus anatomical correlate rather than genuine sinus-region reasoning—a plausible explanation for it being the weakest of the eight vision architectures (Table 3), and a useful reminder that a correct prediction and a correct reason for it are not the same thing.

The language arm makes explicit how much diagnostic information the structured findings carry. Even a shallow model classifies them with exceptionally high accuracy, and architectural diversity makes little difference, indicating that the signal lives in the findings text itself. The ten worked examples in Table 1 show how directly these descriptions encode sinus status. Text-based and image-based performance measure different inputs and should be reported as complementary, modality-specific results rather than pooled into a single headline metric.

Beyond the ten worked examples in Table 1, a broader pattern emerges across the full cohort: the radiologist reclassified 62 of 297 cases (21%) relative to the provisional ChatGPT-only label, almost evenly split between under-calls (32 cases ChatGPT read as normal that the radiologist judged abnormal, chiefly mild mucosal thickening) and over-calls (30 cases ChatGPT read as abnormal that the radiologist judged normal). Table 2 illustrates five of these reclassifications, selected because the radiologist’s correction carried the most additional diagnostic detail rather than a bare label flip. In two cases (images 477.17 and 239.10) ChatGPT reported no abnormal finding at all (“Diagnosis: None” or “no significant paranasal sinus abnormality detected”), yet the radiologist identified moderate mucosal thickening and a retention cyst respectively; a further case (203.12) shows the same pattern for mild mucosal thickening. Conversely, in two cases (364.6 and 4.6) ChatGPT asserted a retention pseudocyst and mucosal thickening that the radiologist did not confirm, reclassifying both as normal. These reclassifications altered only the classification label used for training, not the underlying findings text (Methods), so the pre/post AUC collapse reported for the language and VLM-text arms (Tables 4–6) reflects the label change itself rather than any rewriting of the description; nonetheless, the scale and directionality of these 62 corrections underscore why radiologist-confirmed labels, rather than ChatGPT’s provisional read, are the appropriate reference standard for this benchmark.

The contrastive VLMs reinforce the same lesson under linear probing. CLIP ViT-B/16 and BiomedCLIP recovered only modest image-only discrimination, well below trained convolutional networks, while their text-only probes were exceptionally accurate: any encoder given the radiologist-verified findings text separates the classes almost perfectly. General-purpose VLMs cannot yet be deployed for dental sinus screening on the image alone without task-specific adaptation, and their apparent multimodal competence here derives from the findings text rather than from image understanding.

The generative note-drafting task illustrates a more realistic near-term role for multimodal models: AI-assisted documentation when preliminary findings already exist, not autonomous diagnosis. The key discipline is never to present findings-conditioned generation as independent image interpretation, exactly the distinction the language arm shows are so easily blurred. Even with findings supplied as input, the most verbose model (Qwen2·5-VL) asserted an unsupported laterality in roughly one in nine abnormal notes, whereas LLaVA-Med added none, so model choice and human verification remain essential before any AI-drafted note enters a clinical record.

Several limitations are worth acknowledging: the dataset comprises 300 two-dimensional mid-sagittal slices from a single public source; a single slice cannot capture the three-dimensional extent of sinus disease, and the fractures and soft-tissue involvement central to maxillofacial trauma are inherently volumetric, so performance on full three-dimensional CBCT volumes and on prospectively collected clinical data remains to be established. The reference standard was derived in part from LLM readings before radiologist confirmation, which may introduce systematic bias. The binary normal/abnormal scheme collapses clinically heterogeneous conditions and does not capture severity. The zero-shot VLMs were evaluated with a fixed prompt; lightweight adaptation might improve them. The generative note-drafting arm is exploratory and qualitative rather than formally scored. The generative note-drafting arm is exploratory and qualitative rather than formally scored. This study used the CBCT image slices from the MMDental dataset but did not merge the linked medical-record metadata, including patient sex; the parent dataset reports an approximately balanced sex distribution (51.06% male, 48.94% female), but sex-stratified model performance could not be assessed here. The dataset is drawn from a single hospital in China, and race/ethnicity was not recorded in the source medical records, limiting assessment of generalizability across populations.

On a balanced public dataset, trained convolutional and NAS networks provided a usable image-only baseline for maxillary-sinus CBCT screening (best AUC 0·880), while a ViT underperformed at this sample size. Near-perfect language-model performance reflected the high predictive value of the radiologist-verified findings text rather than image-based skill, a pattern that recurred in the text embeddings of every probed VLM. Multimodal dental AI benchmarks must be interpreted with explicit attention to training regime and data provenance: headline metrics from different modalities are not interchangeable, and transparent, modality-aware reporting is essential to avoid overstating capability. Full three-dimensional CBCT volume analysis is the essential next step for clinical translation in the maxillofacial trauma setting.

## Contributors

SA: methodology, software, formal analysis, investigation, and writing the original draft.

HK: independently reviewed and diagnosed every CBCT image, establishing the radiologist-confirmed reference standard used throughout the study, and review.

TDP: conceptualisation, methodology, supervision, writing, review, and editing. All authors read and approved the final manuscript.

## Data sharing

The MMDental dataset is publicly available.^6^ ChatGPT-generated text and radiologist’s text files are available at the corresponding author’s home page under the heading “ChatGPT generated text and radiologist’s text files”: https://sites.google.com/view/tuan-d-pham/codes. Code used to generate the results is available at https://github.com/sebaalhebshi/sinus-cbct-benchmark (commit 32d5595, 8 August 2026).

## Data Availability

The MMDental dataset is publicly available. ChatGPT-generated text and radiologist's text files are available at the corresponding author's home page under the heading "ChatGPT generated text and radiologist's text files": https://sites.google.com/view/tuan-d-pham/codes. Code used to generate the results is available at https://github.com/sebaalhebshi/sinus-cbct-benchmark (commit 32d5595, 8 August 2026).

https://doi.org/10.6084/m9.figshare.28505276

https://sites.google.com/view/tuan-d-pham/codes

